# Large-scale transcriptome-wide profiling of microRNAs in human placenta and maternal plasma at early to mid gestation

**DOI:** 10.1101/2020.08.19.20177873

**Authors:** Melanie D. Smith, Katherine Pillman, Tanja Jankovic-Karasoulos, Dale McAninch, Qianhui Wan, K. Justinian Bogias, Dylan McCullough, Tina Bianco-Miotto, James Breen, Claire T. Roberts

## Abstract

**Background:** MicroRNAs (miRNAs) are increasingly seen as important regulators of placental development and opportunistic biomarker targets. Given the difficulty in obtaining samples from early gestation and subsequent paucity of the same, investigation of the role of miRNAs in early gestation human placenta has been limited. To address this, we generated miRNA profiles using 96 placentas from presumed normal pregnancies, across early gestation, in combination with matched profiles from maternal plasma. Placenta samples range from 6-23 weeks’ gestation, a time period that includes placenta from the early, relatively low but physiological (6-10 weeks’ gestation) oxygen environment, and later, physiologically normal oxygen environment (11-23 weeks’ gestation).

**Results:** We identified 637 miRNAs with expression in 86 samples (after removing poor quality samples), showing a clear gestational age gradient from 6-23 weeks’ gestation. We identified 374 differentially expressed (DE) miRNAs between placentas from 6-10 weeks’ versus 11-23 weeks’ gestation. We see a clear gestational age group bias in miRNA clusters C19MC, C14MC, miR-17∼92 and paralogs, regions that also include many DE miRNAs. Proportional change in expression of placenta-specific miRNA clusters was reflected in maternal plasma.

**Conclusion:** The presumed introduction of oxygenated maternal blood into the placenta (between ∼10-12 weeks’ gestation) changes the miRNA profile of the chorionic villus, particularly in placenta-specific miRNA clusters. Data presented here comprise a clinically important reference set for studying early placenta development and may underpin the generation of minimally invasive methods for monitoring placental health.

## Background

The human placenta is a complex, rapidly developing and highly regulated organ shared between two genetically unique individuals, the mother and her fetus. Its function is vital for the transfer of nutrients, gases and wastes between maternal and fetal circulations, with dysfunction leading to placental insufficiencies and pregnancy complications [1,2]. Essential placental functions are mediated by the syncytiotrophoblast, large multinucleated cells covering the chorionic villi and providing a large surface area for bi-directional exchange of molecules between fetal and maternal circulations.

Early development of the human placenta occurs in a relatively low, but physiological, oxygen environment due to the occluding presence of extravillous cytotrophoblasts (EVTs) in the uterine spiral arterioles. The placenta transitions to that which is usually considered a physiologically normal, oxygen environment over several weeks from 10 weeks’ gestation when EVTs become dislodged permitting maternal blood to flow into the placental intervillous space [1,3]. This period of rising oxygen tension is a critical time for placental development, as an appropriate response to the accompanying burst of oxidative stress is crucial to the ongoing success of the pregnancy [3]. Robust characterisation of important regulatory mechanisms surrounding normal placental development can give insight into pregnancy health, and the potential development of pregnancy complications [2,4]. Identification of biomarkers of ectopic oxygenation or placental dysfunction leading to pregnancy complications is an important step in monitoring pregnancy health.

MicroRNAs (miRNAs) are an important class of RNA molecules that can help us to understand regulation of developmental pathways within human tissues. As such they are prime targets for biomarker research. miRNAs are short, approximately 20-24 nucleotide (nt) lengths of single-stranded, non-coding ribonucleic acid (RNA) originating in non-coding regions of the genome [5] and comprehensively reviewed in Bartel (2018) [6]. They most commonly function as post-transcriptional regulators of gene expression either by translational repression or targeted degradation by cleavage of mRNA transcripts in the cytoplasm [7,8]. miRNAs target mRNA regulatory sequences that are often located in the 3’ untranslated region (UTR) of the mRNA transcript with potential for multiple conserved target sites within a single mRNA [5,9]. Approximately half of known mammalian miRNAs are found within intergenic regions of the genome, with the remaining miRNAs mostly transcribed from the open reading frame of other genes, including long non-coding RNA (lncRNA) [10–12]. Adding to this complexity, miRNA may be transcribed as a single, or polycistronic unit [11].

The critical role of the placenta in successful pregnancy is clear and there is also evidence linking developmental programming of chronic adult disease such as heart disease, diabetes and obesity with the placental phenotype *in utero* [4,13,14]. However, there are currently no non-invasive tests to determine which women are likely to develop pregnancy complications in routine use, nor is there a comprehensive reference set of miRNA expression during early pregnancy that can be used for development of pregnancy health biomarkers. In this study, we performed high-throughput sequencing to generate robust miRNA expression profiles for 96 placentas and matched maternal plasma samples between 6-23 weeks’ gestation, creating a comprehensive set of miRNA expression profiles for presumably healthy placental function. We identified differential miRNA expression between 6-10 and 11-23 weeks’ gestation which could reflect physiological changes occurring in this important time period. Using placenta-associated miRNA clusters found on chromosome 14, 19, 13, 7 and X, we also investigated how maternal plasma reflects placental profiles to enable future biomarker discoveries.

## Results

Characterising a comprehensive miRNA profile of chorionic villous tissue across early gestation to characterise the human placental miRNA profile across early to mid gestation, Illumina NextSeq 75 bp single-end read sequencing was performed on miRNA libraries from chorionic villous samples obtained from 96 singleton pregnancies following elective pregnancy terminations (44 female and 52 male bearing pregnancies). An average of 18.72 million reads were sequenced per sample (range ∼10-29 million reads; Additional file 1: Table S1). miRNA counts were generated by mapping to human miRBase V21 [15]. Libraries with low sequencing depth were removed, including samples PAC0131 (14,595 reads) and PAC0071 (removed due to low mapping after initial read count of 1,819,847 reads). Subsequent unsupervised clustering (Additional file 2: Figure S1) using principal component analysis (PCA) showed eight additional samples (PAC0041, PAC0039, PAC0045, PAC0008, PAC0035, PAC0024, PAC0034, PAC0006) that segregated from the remaining 86 samples, most likely due to the presence of adjacent non placental villous tissue. This hypothesis was tested using matched DNA methylation profiles which confirmed the presence of decidual tissue [16], leading to removal of these samples from subsequent analyses. After filtering, miRNAs were aligned to the human GRCh37 reference genome and counts produced from 4,665 miRBase annotations [15], identifying an initial set of 2,032 expressed miRNAs. To remove sequencing noise, miRNAs with <5 reads across all samples were removed. A total of 1,422 miRNAs were identified with non-zero expression means across all 86 samples, representing a standard known miRBase miRNA expression set for early to mid gestation human placenta. After accounting for sequencing batch effects, very low abundance miRNAs were discarded by converting to log_2_ CPM values and removing miRNAs with expression <2.5 CPM in ≥29 samples (see methods), leaving a total of 637 robust miRNAs for downstream profile analyses. An additional 588 candidate *de novo* miRNA sequences were predicted through miRDeep2 analysis.

Of the 637 identified miRNAs, the top ten miRNAs with the highest mean expression across all samples in placenta were miR-30d-5p, miR-125a-5p, miR-517a-3p, miR-199a-3p, miR-26b-5p, miR-26a-5p, let-7a-5p, miR-21-5p, miR-126-3p and miR-516b-5p (Table 1a). For a full list of miRNAs identified and profiled in this study see Additional File 1: Table S2. A principal component analysis (PCA) of all 637 miRNAs (Figure 1) indicates a clear gradient across early gestation. The PCA identified 23.2% variance accounted for in PC1, with the top 10 miRNAs contributing to PCA dimension one (PC1) being: miR-519c-3p, miR-328-3p, miR-519b-3p, miR-20a-5p, miR-19b-3p, miR-501-3p, miR-485-5p, miR-485-5p, miR-3605-3p and miR-515-5p (Additional file 1: Table S3). Interestingly both -3p and -5p arms of the miR-20a transcript, previously associated with placental angiogenesis [17], contributed highly to the gestational age change associated with PC1.

**Table 1:** miRNA with highest expression in early gestation placenta. Global miRNA expression was calculated using the average expression across early gestation (6-23 weeks’) and log_2_ transformed. (**a**) Top ten miRNA with highest average global expression. (**b**) Constitutively highly expressed miRNA with membership in the top ten by highest average expression if calculated independently for the 6-10 weeks’ and 11-23 weeks’ gestation groups.

| a |  |  | b |  |  |  |
| --- | --- | --- | --- | --- | --- | --- |
| miRNA | miRNA Family | Avg. Expression ($\log_2$ CPM) | miRNA | miRNA Family | Avg. Expression ($\log_2$ CPM) | |
|  |  |  |  |  | 6-10 weeks' | 11-23 weeks' |
| miR-30d-5p | miR-30-5p | 15.60 | miR-30d-5p | miR-30d-5p | 15.53 | 15.64 |
| miR-125a-5p | miR-125-5p | 15.51 | miR-125a-5p | miR-125-5p | 14.93 | 15.74 |
| miR-517a-3p | miR-517-5p | 15.45 | miR-517a-3p | miR-517-5p | 15.52 | 15.40 |
| miR-199a-3p | miR-199-3p | 15.43 | miR-199a-3p | miR-199-3p | 15.21 | 15.56 |
| miR-26b-5p | miR-26-5p | 15.25 | miR-26b-5p | miR-26-5p^ | 15.27 | 15.21 |
| miR-26a-5p | miR-26-5p | 14.84 | let-7a-5p | let-7-5p/98-5p | 13.59 | 14.83 |
| let-7a-5p | let-7-5p/98-5p | 14.47 | miR-21-5p | miR-21-5p/590-5p | 14.20 | 14.59 |
| miR-21-5p | miR-21-5p/590-5p | 14.39 | miR-126-3p | None listed | 13.91 | 14.56 |
| miR-126-3p | None listed | 14.31 | miR-516b-5p | miR-516b-5p | 13.23 | 14.31 |
| miR-516b-5p | miR-516b-5p | 14.30 | miR-16-5p | miR-15-5p/16-5p/195-5p/424-5p/497-5p | 14.34 | 14.32 |
|  |  |  | miR-516a-5p | miR-516a-5p | 14.40 | 14.11 |
|  |  |  | miR-143-3p | miR-143-3p | 13.60 | 14.52 |
|  |  |  | miR-26a-5p | miR-26-5p^ | 14.54 | 14.99 |

^shared seed region

**Figure 1:**
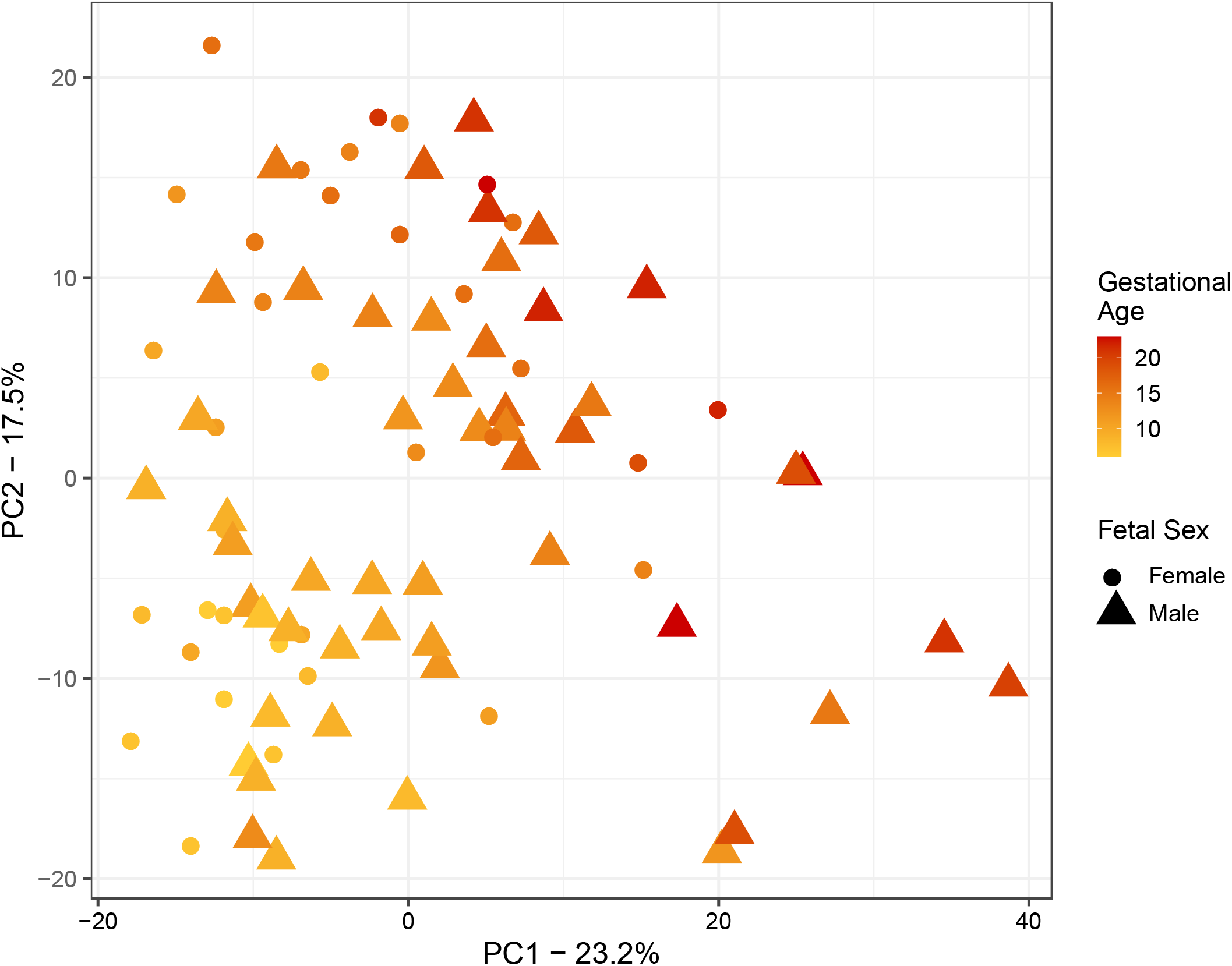
Unsupervised clustering of placental chorionic villous samples after batch correction. The PCA plot indicates a clear gestational age gradient from early (left) to later gestation (right) in the first dimension. This trend reflects the gestational age signature identified in our miRNA expression data.

### Differentially expressed miRNAs across early gestation

Given the importance of oxygen tension in orchestrating the growth and differentiation of the placenta between 6–23 weeks’ gestation, we separated the 86 samples in our cohort by their gestation relative to the presumed introduction of oxygenated maternal blood flow into the intervillous space. Using prior knowledge we designated that ≤10 weeks’ (6-10 weeks’) gestation be considered pre- and >10 weeks’ (11-23 weeks’) gestation be considered post-initiation of maternal blood flow into the intervillous space [3]. This enabled us to identify changes in miRNA expression across this time-point, and to independently assess highly expressed miRNA in each group. When miRNAs from all 86 samples were analysed according to expression prior to and post 10 weeks’ gestation, after accounting for potentially confounding factors such as maternal age, maternal BMI, maternal smoking status, we identified 13 constitutively expressed miRNAs with consistent high expression (log_2_ CPM 14.35 - 15.60) in both gestation groups (Table 1b). Notably, four of the top twenty most highly expressed miRNAs are from the placenta-specific chromosome 19 cluster, previously identified as being highly expressed in placenta [18–20].

After identifying the highly expressed miRNAs in placenta, we then investigated the change in expression that occurs after 10 weeks’ gestation once maternal blood flow into the placenta is initiated and presumably when oxygen tension begins to rise. Differential expression analysis identified 374 significantly (FDR <0.05) different miRNAs, with 163 down-regulated and 211 up-regulated in the 11-23 weeks’ gestation placenta compared to 6-10 weeks’ gestation (Figure 2a; Additional file 1: Table S4). Strongly downregulated miRNAs (negative log_2_FC) include miR-4483, miR-129-5p, miR-124-3p and miR-122-5p, and highest up-regulated (positive log_2_FC) include miR-4645-3p, miR-195-3p, miR-137, miR-139-3p, miR-6715b-3p, miR-3927-3p and miR-1269b. Differentially expressed miRNAs had predominantly low expression (Figure 2b) with an average log_2_ expression >10 only observed in miR-9-5p (log_2_FC of −1.43, Figure 2c) and let-7a-5p (log_2_FC of 1.22, Figure 2d). We see that for both let-7 and miR-9, the 5’ and 3’ arms follow a similar expression trajectory. These mature fragments were originally transcribed as a single transcript before further processing (Figure 2e).

**Figure 2:**
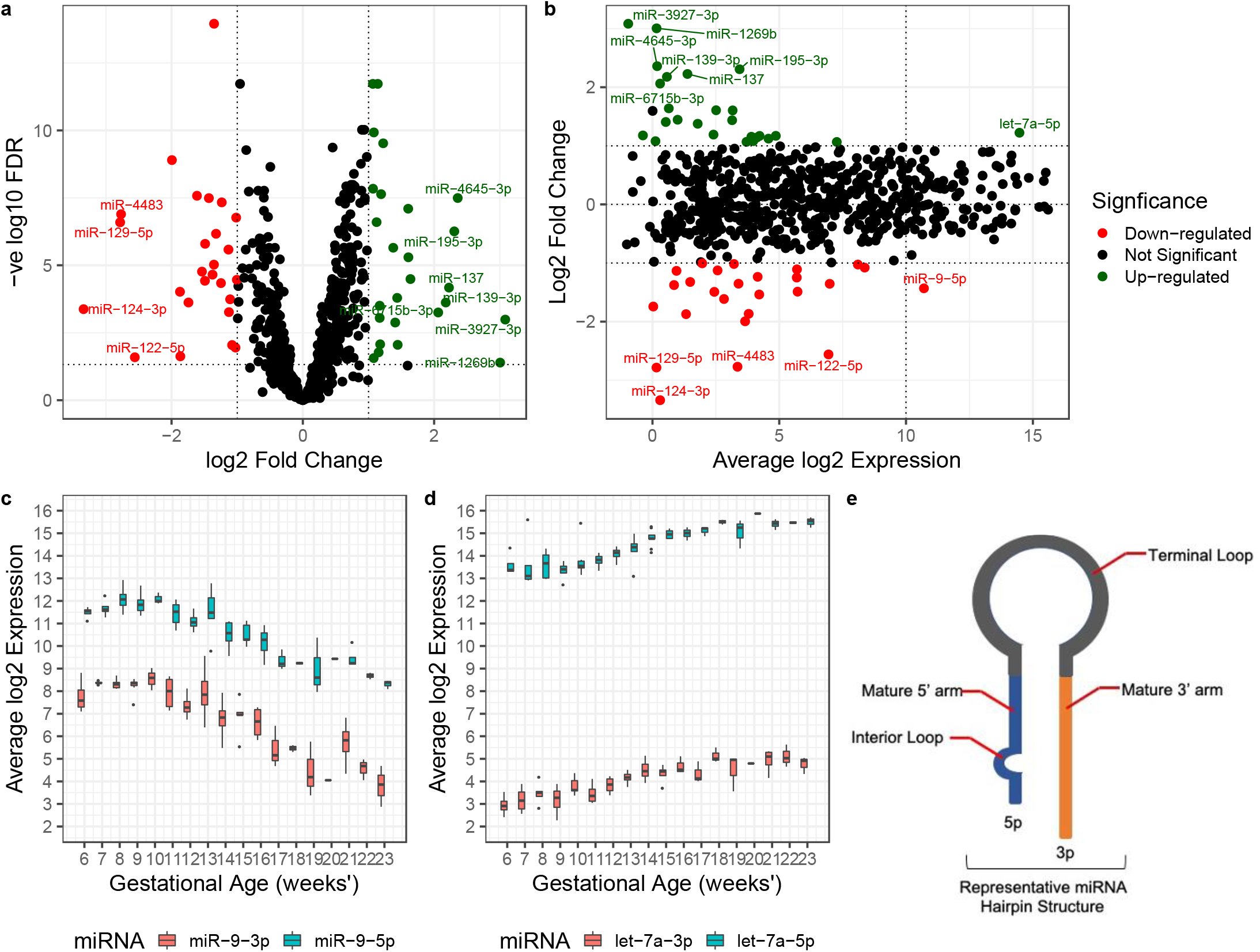
Differential expression analysis between 6-10 weeks’ and 11-23 weeks’ gestation placenta. (**a**) Volcano plot of differential expression. Linear regression identified 26 down-regulated with a log_2_FC <-1 (red) and 26 up-regulated with a log_2_FC >1 (green) miRNA between gestation groups. (**b**) MA plot of log_2_ fold change as a function of log_2_ average expression. Box plots of average expression as a function of gestational age for miR-9-5p/3p (**c**) and let-7a-5p/3p (**d**) show a clear changing pattern of expression from early to mid gestation. **(e)** Schematic of a representative miRNA hairpin structure indicates the origin of the 5’ and 3’ arms of the mature miRNA transcript.

### Conserved placenta-associated miRNA clusters display variable expression between early and mid gestation

In vertebrate species miRNAs are significantly enriched in clusters, with ∼50% of miRNA clusters being the result of random or non-local duplications. The placenta-associated clusters analysed herein included chromosome 19 miRNA cluster commonly known as C19MC (19q13.41) [21], chromosome 14 cluster (C14MC; 14q.32) [22], and chromosome 13 cluster (miR-17∼92; 13q31.3) with its paralogs the chromosome X cluster miR-106a∼363 (Xq26.2), and chromosome 7 cluster miR-106b∼25 (7q22.1) [23]. We identified the expression of 48 mature miRNAs from C19MC, 77 from C14MC and 20 from miR-17∼92 and paralogs. Of the 48 mature miRNAs identified in the C19MC cluster, 34 miRNA had significant (FDR <0.05) differential expression, that were down regulated in 11-23 weeks’ compared to 6-10 weeks’ gestation placenta (Additional file 1: Table S5). C19MC cluster expression, as a proportion of total miRNA transcripts, was reduced after 10 weeks’ gestation (Figure 3a). Interestingly, while miRNAs in this cluster have been demonstrated to be transcribed by RNA-Pol II as a single polycistron [24], the individual members display a range of expression levels (Figure 3b).

**Figure 3:**
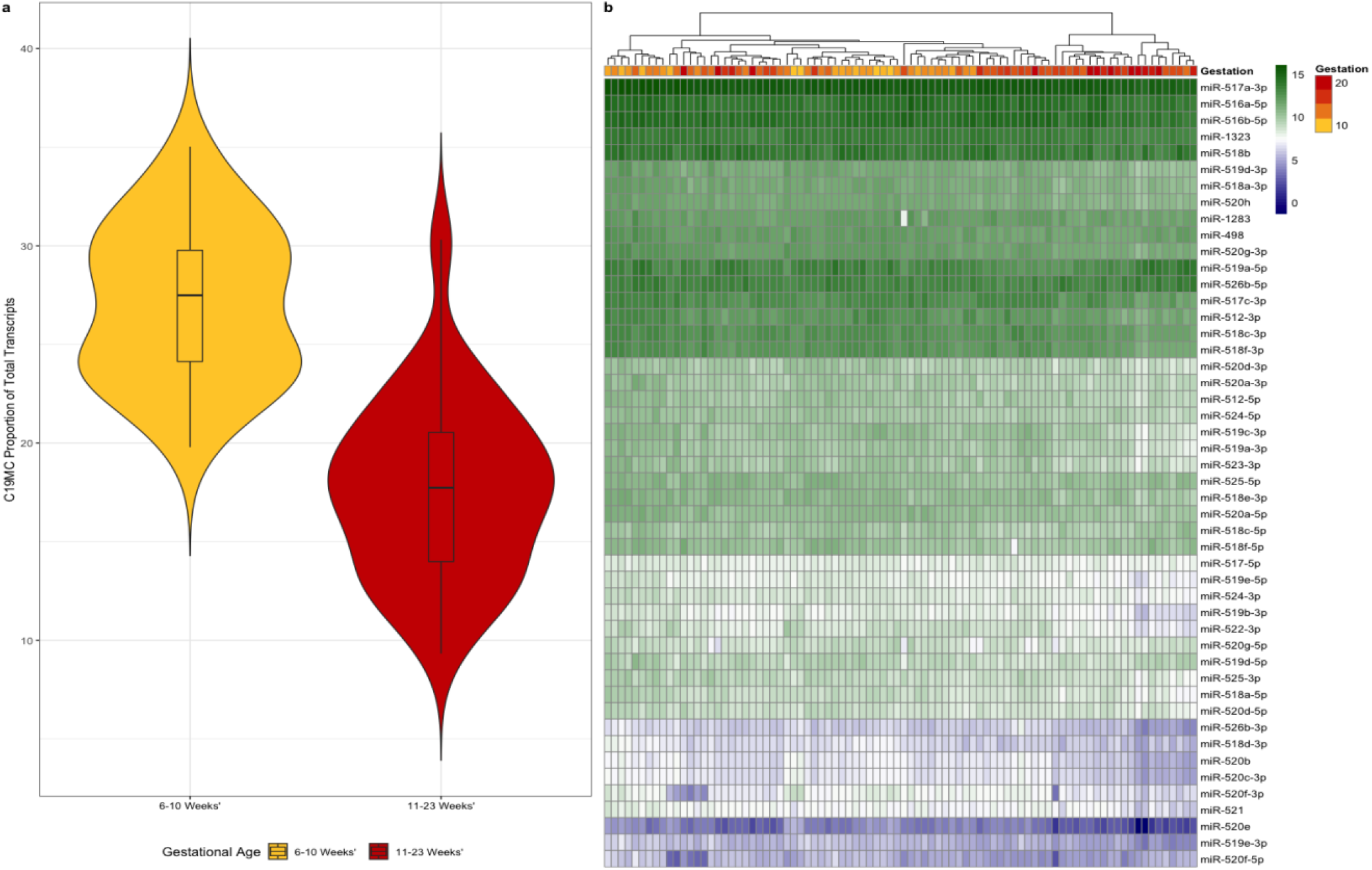
The proportion of C19MC transcripts decreased after 10 weeks’ gestation in placenta. The proportion of combined C19MC member expression as a function of total miRNA expression is decreased in 11-23 weeks’ compared to 6-10 weeks’ gestation (violin plot) (**a**) with expression of individual cluster members tending to decrease across early to mid gestation (**b**).

The C14MC cluster, that contains 77 mature miRNAs, had 56 miRNAs with significant (FDR <0.05) differential expression. 55 of these miRNAs were up-regulated, while only one miRNA (miR-410-5p; FDR 1.99 x 10^×3^) was down-regulated in the 11-23 weeks’ gestation group (Additional file 1: Table S6). C14MC-member transcript expression, as a proportion of total expression, increased across early to mid gestation in placenta, further differing from C19MC finding by being more centered around the group mean (Figure 4a). Similar to C19MC cluster members, we found a range of expression levels amongst the C14MC members (Figure 4b) that are also thought to be transcribed as a single polycistronic transcript, suggesting more complex regulation of primary miRNA transcripts processing [25]. The miRNA miR-412-5p, previously observed as highly expressed in first trimester placenta [26], is notable in that it appears to have a bimodal distribution of expression (Figure 4c) that, upon further investigation, was not found to be related to sample characteristics such as fetal sex, maternal age, maternal BMI or maternal smoking status.

**Figure 4:**
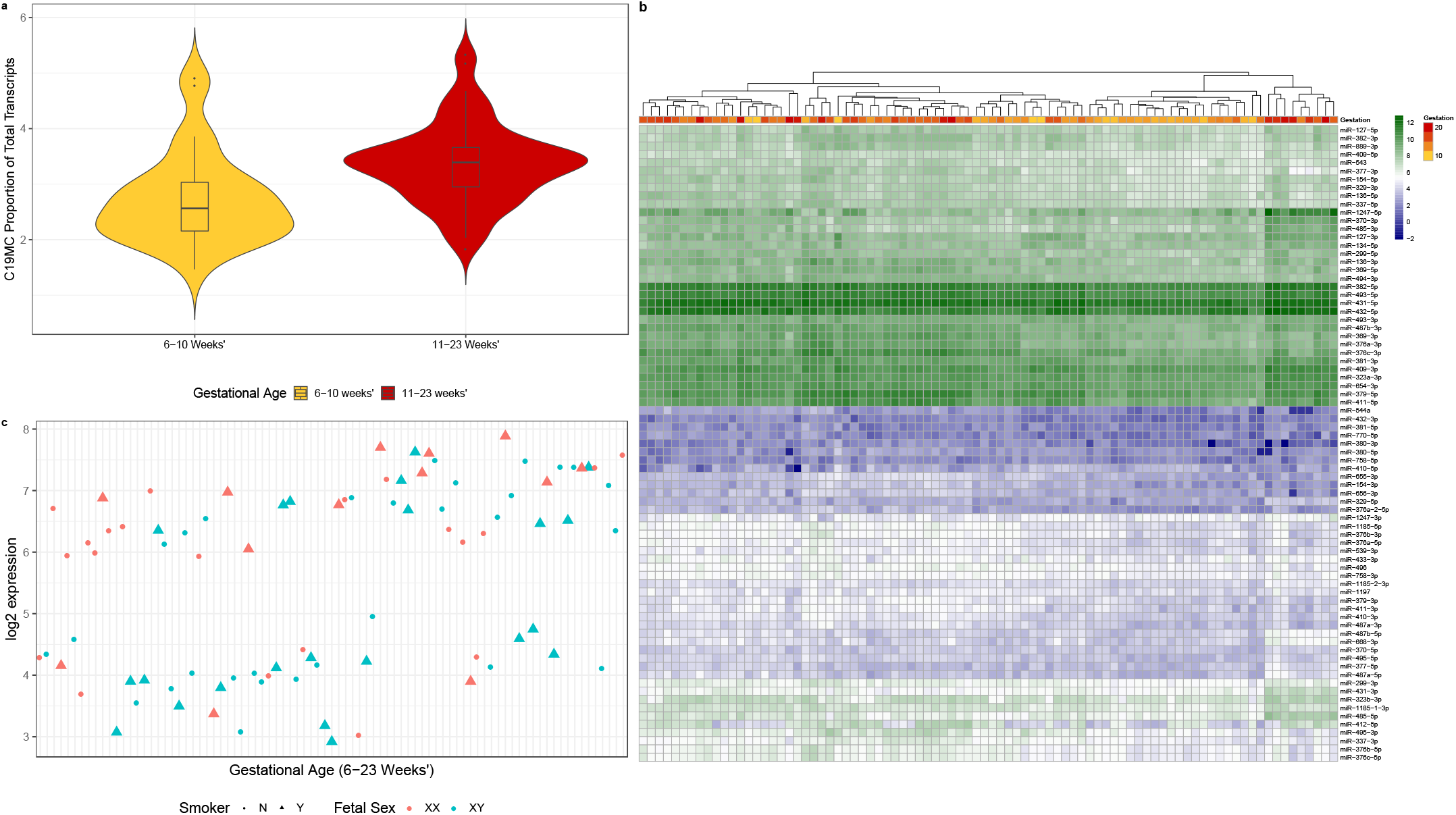
The proportion of C14MC transcripts increased in 11-23 weeks’ compared to 6-10 weeks’ gestation placenta. The proportion of C14MC member expression as a function of total miRNA expression is increased after 10 weeks’ gestation compared to 6-10 weeks’ gestation (violin plot) (**a**). Individual cluster members’ expression is increased across early gestation in 55 of the 56 miRNAs (**b**). miR-412-5p appears to have a bimodal distribution which is unexplained by fetal sex, maternal age or maternal smoking status (**c**).

Of the 20 mature miRNAs in the miR-17∼92 cluster and its paralogs, 17 had significant (FDR <0.05) differential expression with 15 being down- and 2 being up-regulated in the 11-23 weeks’ gestation group compared to the 6-10 weeks’ (Additional file 1: Table S7). Investigation into the proportion of placenta expression originating from the miR-17∼92 cluster and its paralogs revealed a pattern similar to that of the C19MC member transcripts with the proportion of transcripts identified in placenta tissue as a function of total transcripts decreasing across early gestation (Figure 5a, b, c). We identified a variation in expression patterns between each of the three paralogs (Figure 5d), with chromosome 13 expression decreasing across early to mid gestation.

**Figure 5:**
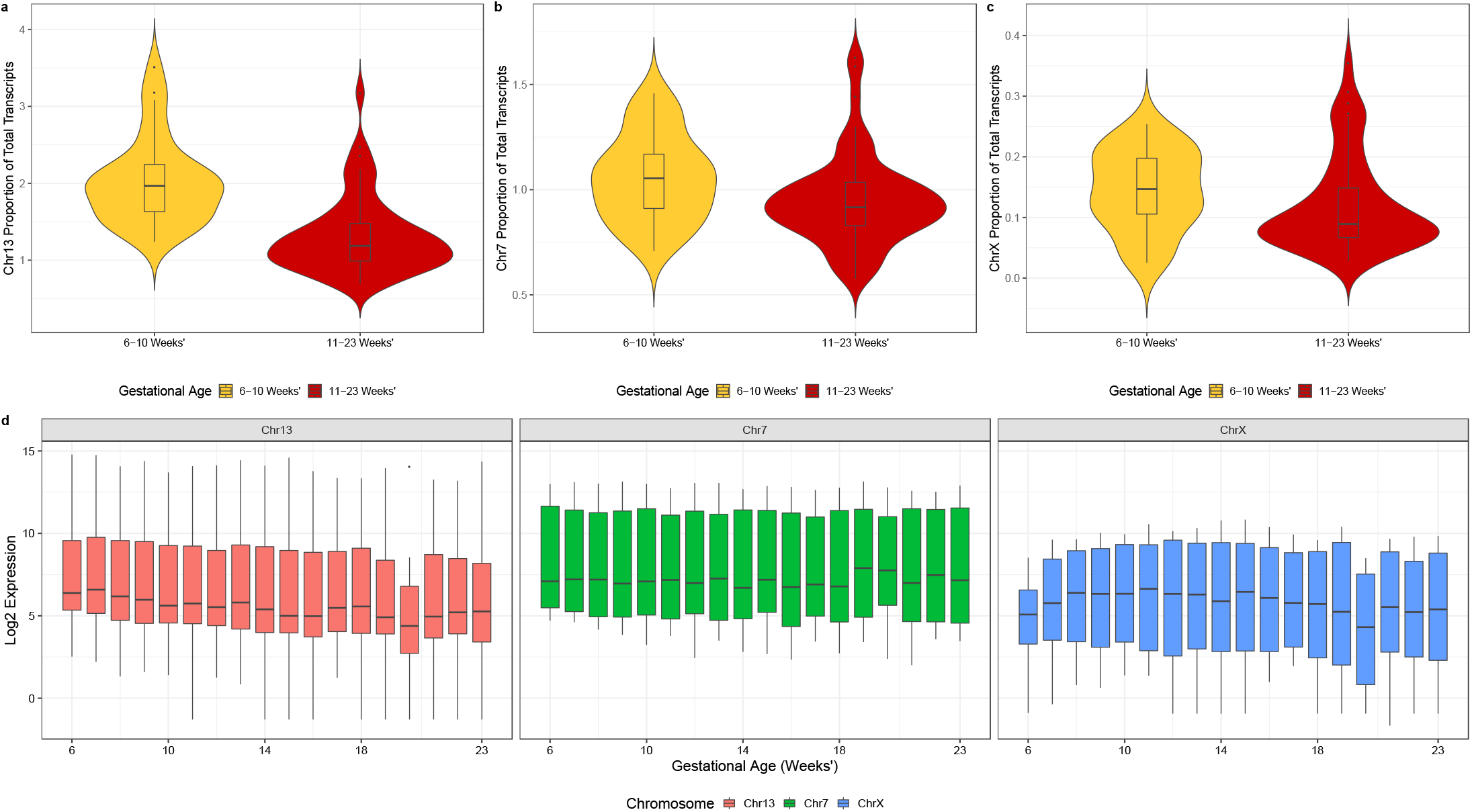
The proportion of miR-17∼92 (Chr 13) and paralogs 106b∼25 (Chr 7) and 106a∼363 (Chr X) transcripts decreased in 11-23 weeks’ compared to 6-10 weeks’ gestation placenta. Violin plot showing decreased proportion of miR-17∼92 member expression as a function of total miRNA expression after 10 weeks’ gestation (**a, b, c**) with a general trend of decreased expression of individual cluster members across early gestation (**d**). Although chromosome 13 and chromosome X appear correlated, expression of miR-17∼92 (chromosome 13) and its paralogs miR-106a∼363 (chromosome 7) and miR-106b∼25 (X chromosome) does not appear to follow a common expression pattern in placental tissue as we would expect if all 3 clusters are identically regulated by C-Myc.

Whilst we find significant (FDR <0.05) differential expression in all of the above miRNA clusters, the log_2_FC between 6-10 weeks’ and 11-23 weeks’ gestation groups is small (C19MC log_2_FC −1.02 to −0.19; C14MC log_2_FC 0.23 to 0.94 and log_2_FC 0.67; miR-17∼92 log_2_FC −0.2 to −1.13 and log_2_FC 0.84 to 0.89), with changes in the proportional expression of each cluster as a function of total miRNA expression suggesting an important regulatory mechanism for these clusters in placenta.

### Characterising a comprehensive profile of placental and endogenous miRNA in maternal plasma across early gestation

To characterise the miRNA profile across early to mid gestation in maternal plasma, we sequenced 96 matched maternal plasma samples (hereafter referred to as plasma) from the same women sampled at the time of pregnancy termination using the same sequencing protocol as detailed for the chorionic villous tissue. miRNA sequencing resulted in approximately 17.77 million 75 bp single-end reads per sample in plasma (range 9,986,347-41,344,361 reads: Additional file 3: Table S8) across two sequencing runs. After initial quality control and sequence analysis, we identified a number of samples with aberrant expression of miR-451a and miR-16 indicating potential for red blood cell miRNA contamination. We performed a subsequent analysis using the delta-delta CT method (ΔΔCt) to calculate the relative fold change of gene expression of samples using quantitative polymerase chain reaction (qPCR). The ΔΔCt value of miR-23a minus miR-451 is examined, as miR-23a is known to be stable in samples affected by haemolysis whilst miR-451 is known to vary with contamination. This method identified 11 samples (PAC0033, PAC0034, PAC0041, PAC0048, PAC0050, PAC0051, PAC0054, PAC0056, PAC0062, PAC0084 and PAC0105) as more likely to contain haemolysis contamination, a common issue in plasma research. Despite potential issues involving haemolysis, we aimed to establish the presence of placenta specific miRNAs within maternal plasma by including all available samples regardless of whether they had matched placenta data or not.

Alignment and filtering were performed using the same protocol as for chorionic villous tissue. The number of unique miRNA species identified per sample varied, ranging from 1022 miRNA species in PAC0077 to 355 miRNA species in PAC0010. Prior to filtering we identified 1775 unique mature miRNAs across all plasma samples, with a core set of 790 high confidence miRNAs present after filtering. Of these, 531 were also present in placenta samples, 259 unique to plasma, and 106 miRNAs unique to placenta (Additional file 3: Table S9). Unsupervised clustering (PCA) of all 790 high confidence miRNAs identified in plasma shows no gestational age gradient for maternal plasma from 6-23 weeks’ gestation (Additional file 2: Figure S2).

Given the variability of miRNA in plasma, we hypothesised that detection of placental miRNAs in plasma may be restricted to miRNA present in high concentrations in matched placenta. We therefore investigated whether ten miRNAs (Table 1a) with the highest abundance in placenta were present in plasma and found that all ten miRNA were at high abundance levels in matched plasma (Figure 6, Table 2a). Many of the most highly expressed plasma miRNA are red blood cell (RBC) associated.

**Table 2:** miRNA by average expression in maternal plasma. Average miRNA expression was calculated using the average normalised CPM expression across early to mid gestation (6-23 weeks’) and log_2_ transformed. **(a)** All top 10 miRNA with highest abundance in placenta were also detected at high abundance in plasma. Two miRNA (miR-517a-3p and miR-516b-5p) originate from the C19MC. **(b**) Top 10 miRNA by abundance in maternal plasma. RBC-associated miRNA identified herein have been previously identified in plasma samples including adult plasma and umbilical cord plasma but have not previously been profiled extensively in maternal plasma in early to mid gestation.

| a |  |  | b |  |  |
| --- | --- | --- | --- | --- | --- |
| miRNA | miRNA Family | Avg. Expression (log<sub>2</sub> CPM) | miRNA | miRNA Family | Avg. Expression (log<sub>2</sub> CPM) |
| miR-30d-5p | miR-30-5p | 13.35 | miR-16-5p<sup>+</sup> | miR-15-5p/16-5p/195-5p/424-5p/497-5p | 18.23 |
| miR-125a-5p | miR-125-5p | 10.59 | miR-486-5p<sup>+</sup> | miR-486-5p | 17.46 |
| miR-517a-3p<sup>^</sup> | miR-517-5p | 4.71 | miR-92a-3p<sup>+</sup> | miR-25-3p/32-5p/92-3p/363-3p/367-3p | 16.22 |
| miR-199a-3p | miR-199-3p | 10.62 | let-7a-5p | let-7-5p/98-5p | 15.70 |
| miR-26b-5p | miR-26-5p | 12.84 | let-7b-5p | let-7-5p/98-5p | 15.55 |
| miR-26a-5p | miR-26-5p | 12.70 | miR-451a<sup>+</sup> | miR-451 | 14.42 |
| let-7a-5p | let-7-5p/98-5p | 15.70 | let-7f-5p | let-7-5p/98-5p | 14.15 |
| miR-21-5p | miR-21-5p/590-5p | 12.87 | miR-25-3p | miR-25-3p/32-5p/92-3p/363-3p/367-3p | 13.97 |
| miR-126-3p | None listed | 13.41 | let-7i-5p | let-7-5p/98-5p | 13.92 |
| miR-516b-5p<sup>^</sup> | miR-516b-5p | 4.27 | miR-93-5p | miR-17-5p/20-5p/93-5p/106-5p/519-3p | 13.58 |

<sup>^</sup> C19MC
<sup>+</sup> RBC

**Figure 6:**
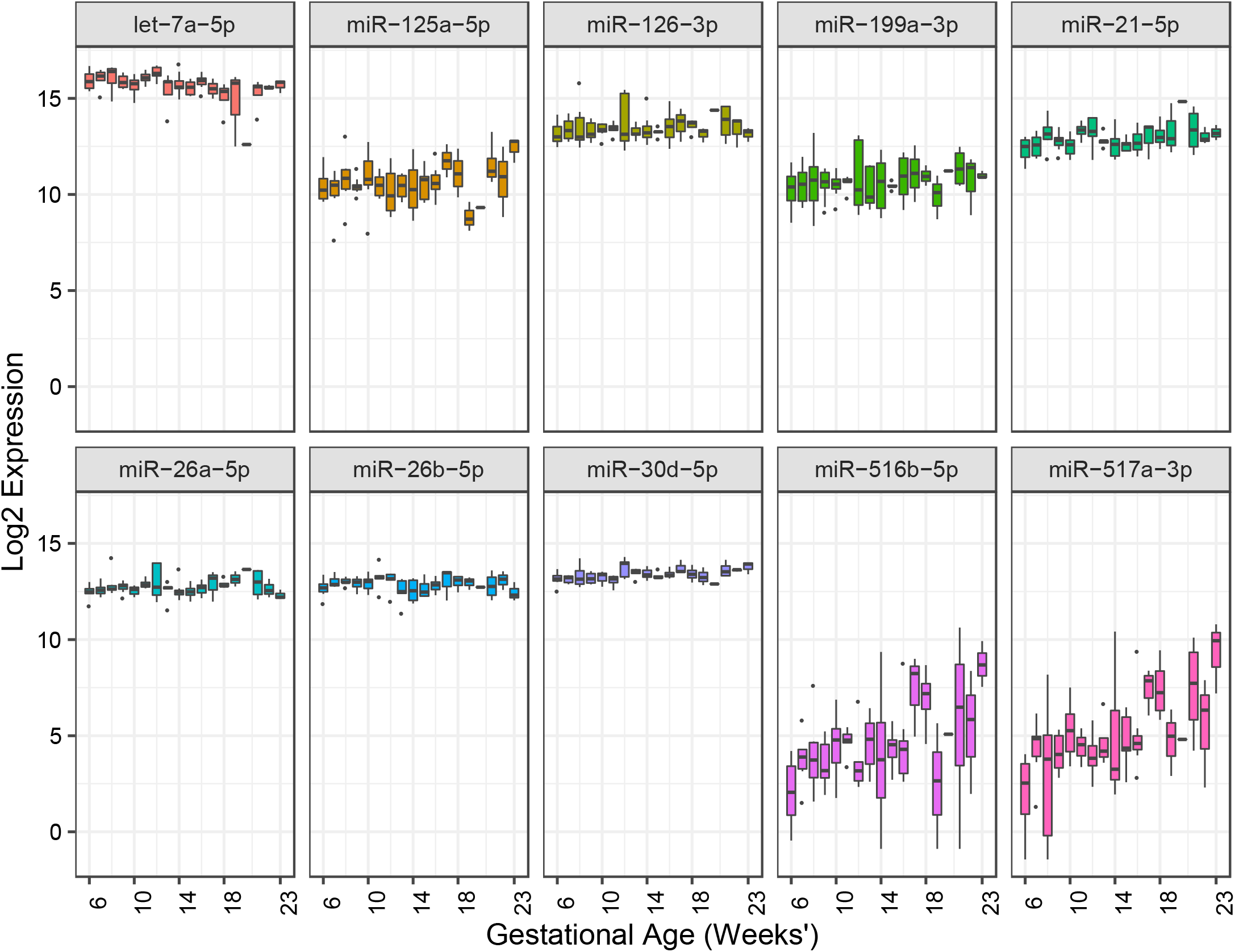
Abundance of maternal plasma miRNAs, identified as highly expressed in chorionic villous tissue, from 6-23 weeks’ gestation. All ten miRNAs identified as the most highly expressed in chorionic villous tissue were identified as highly abundant in matched plasma samples. Abundance of the C19MC miRNA members, miR-516b-5p and miR-517a-3p, appear to increase across early to mid gestation but these were not statistically significant (FDR <0.05).

Of the 790 miRNAs identified in plasma, the top ten miRNAs with the highest global expression were miR-16-5p, miR-486-5p, miR-92a-3p, let-7a-5p, let-7b-5p, miR-451a, let-7f-5p, miR-25-3p, let-7i-5p, miR-93-5p (Table 2b). These were also all highly expressed in placental tissue (average global expression of 8.95-14.47 Log_2_ normalised CPM). miR-141, miR-200b, miR-139 and miR-184, previously identified in third trimester plasma, were also identified at high abundance in our early gestation plasma, with miR-141-3p abundance increasing across early gestation, as shown in previous studies [27].

We investigated plasma levels of the 21 miRNAs identified as both differentially and highly expressed in the matched placentas (Table 3). Whilst all 21 placental miRNAs were also detected in plasma, their origin is not necessarily placenta-specific, with cellular enrichment linked to various systemic compartments [28]. For example, five of these miRNAs have cellular enrichment in immune response, another three miRNAs in pluripotent and embryonic stem cells, and one miRNA is enriched in extra-embryonic stem cells. The latter may originate from the fetal membranes or other extra-embryonic tissue such as chorionic villi.

**Table 3:** The abundance in plasma of miRNA differentially expressed in matched placenta. Average miRNA expression was calculated using the average CPM expression across early to mid gestation (6-23 weeks’) and log_2_ transformed.

| Placenta |  |  | Maternal Plasma |  |  |
| --- | --- | --- | --- | --- | --- |
| Placenta DE miRNA | log<sub>2</sub>FC | AveExp (log<sub>2</sub>) | AveExp (log<sub>2</sub>) | Compartment of origin* | Cellular enrichment^ |
| let-7a-5p | 1.24 | 14.27 | 15.70 | brain, epididymis, spinal cord | Mesodermal cells |
| miR-542-5p | 1.11 | 7.07 | 0.66 | epididymis, small intestine, lymph node | None listed |
| miR-222-3p | 1.00 | 9.66 | 6.51 | bladder, epididymis, prostate | Endothelial cell, endothelial cell of vascular tree, blood vessel endothelial |
| miR-1-3p | 1.00 | 7.20 | 4.45 | muscle, myocardium | Muscle, skeletal muscle, mesodermal cell |
| miR-199b-5p | 0.97 | 10.24 | 2.72 | colon, epididymis, esophagus, skin, thyroid, vein | Fibroblast, mesodermal cell |
| miR-99a-5p | 0.98 | 8.57 | 5.92 | Arachnoid mater, brain, epididymis, prostate, spinal cord | Mast cell, leukocyte, myeloid leukocyte |
| miR-223-3p | 0.99 | 10.93 | 13.28 | vein | Leukocyte, hematopoietic cell, myeloid leukocyte |
| miR-221-3p | 0.94 | 13.13 | 9.65 | Epididymis, prostate | Endothelial cell, ciliated epithelial cell, muscle cell |
| miR-100-5p | 0.98 | 12.83 | 5.58 | Arachnoid mater, nerve, spinal cord, brain | Muscle cell, smooth muscle cell, vascular associated smooth muscle cell |
| miR-181a-5p | 0.94 | 13.06 | 10.09 | Arachnoid mater, brain, dura mater, spinal cord | Leukocyte, hematopoietic cell, B cell |
| miR-363-3p | 0.94 | 9.18 | 9.26 | Epididymis, vein | Leukocyte, hematopoietic cell, lymphocyte |
| miR-122-5p | -2.53 | 6.74 | 13.09 | Liver, vein | Hepatocyte, endodermal cell |
| miR-9-5p | -1.42 | 10.51 | 2.19 | Brain, spinal cord | Neural cell, neuroectodermal cell, ectodermal cell |
| miR-9-3p | -1.37 | 6.79 | -2.25 | Brain, dura mater, spinal cord | None listed |
| miR-372-3p | -1.08 | 8.17 | -2.98 | No profile | Pluripotent stem cell, embryonic stem cell, induced pluripotent stem cell, depleted in extraembryonic stem cell |
| miR-371a-5p | -1.01 | 7.90 | -2.81 | Constitutive expression | Pluripotent stem cell, embryonic stem cell, induced pluripotent stem cell, depleted in extraembryonic stem cell |
| miR-520f-3p | -0.98 | 6.86 | -3.07 | spleen | Pluripotent stem cell, embryonic stem cell, induced pluripotent stem cell, depleted in extraembryonic stem cell |
| miR-934 | -0.93 | 9.32 | -2.44 | Constitutive expression, highest in gallbladder and pleura | Epithelial cell, extraembryonic cell, endo-epithelial cell |
| miR-20a-5p | -0.85 | 10.02 | 9.73 | Constitutive expression, highest in vein | Pluripotent stem cell, epithelial cell, endothelial cell |
| miR-17-5p | -0.77 | 6.80 | 8.77 | Constitutive expression, highest in vein | Epithelial cell, pluripotent stem cell, endothelial cell |
| miR-373-3p | -0.68 | 6.74 | -2.57 | kidney | Pluripotent stem cell, embryonic stem cell, induced pluripotent stem cell |

\* [29]
^ [30]

We further investigated the abundance of plasma miRNAs from three miRNA clusters previously identified as highly placenta specific or placenta associated [21–23]. We found 44 mature miRNA from the C19MC cluster, 48 miRNA from the C14MC cluster and 20 miRNA from the miR-17∼92 cluster and its paralogs in plasma. These cluster miRNAs have previously been the target of biomarker research in pregnancy health but to our knowledge these are not being used clinically.

The change in relative proportion of expression by miRNA cluster members in plasma across the 10-11 weeks’ gestation threshold is difficult to detect with C14MC and C19MC members each representing less than 1% of the miRNA abundance quantified in plasma. As such the proportional change identified in placenta is not entirely reflected in plasma. Abundance levels of the miR-17∼92 cluster and its paralogs represent approximately 0.03% of the total miRNA identified in plasma.

## Discussion

In this study we have assessed the miRNA profile of 96 human placenta and matched plasma samples in early to mid gestation (6-23 weeks’), generating a placental tissue dataset using high-throughput sequencing that is substantially larger than previous reports that used microarray and qPCR profiling. We derive robust and comprehensive profiles that define miRNA expression at high-resolution, applying stringent bioinformatic protocols for the analysis of sequencing data, akin to large atlas studies.

### Differentially expressed miRNAs reflect the dynamically changing utero-placental environment during early to mid gestation

In order to determine the potential association between placental miRNA expression and pregnancy health, particularly the expression of highly abundant miRNAs across early gestation, we examined more closely a period in early gestation involving a steep rise in oxygen tension in the intervillous space (IVS). Rising oxygen tension occurs between 10-12 weeks’ gestation, at onset of maternal blood flow into the IVS, resulting in an oxidative burst in placenta [3]. Previous studies have addressed miRNA response to hypoxia and the introduction of oxygen [31–35], including how ectopic oxygen tension may contribute to diseases of pregnancy [31,36–38]; and reviewed in [39]. The number and extensive gestation range of our samples enabled us to analyse the differential expression of miRNAs prior to, during and post the critical 10-11 weeks’ gestational time point. This has potential to lead to identification of placental mechanisms for the remediation of oxidative stress at this crucial time in pregnancy.

That the placenta develops initially in a condition of low (but physiological) oxygen tension has been known for some time [3] and it has been suggested that this is important for the protection of the early fetus from the effects of oxidative stress such as tissue damage from oxygen free radicals [40]. miRNAs are uniquely poised to play a significant role in the mediation of gene expression throughout the early change of oxygen tension in the placenta by employing post transcriptional regulatory mechanisms to rapidly and precisely fine tune the placental environment in response to oxidative stress. We identified upregulation of let-7b/e5p, miR-21, miR-23a/b-3p, miR-24-3p and miR-199a-5p, also associated with *HIF1α*, in the 6-10 weeks’ gestation group. All of these have previously been shown to be upregulated by hypoxia. We also found lower abundance of let-7a and miR-101-3p in our 6-10 weeks’ gestation group, both of which have previously been shown to be downregulated by hypoxia [41–45]; and reviewed in [46]. miRNAs identified in this study likely play an important role in the mitigation and regulation of hypoxia in the placenta. This may prove important to our understanding of diseases of pregnancy such as preeclampsia and early pregnancy loss.

The importance of miRNA function in the early placenta is not limited to the mitigation of oxidative stress but also in cellular proliferation, trophoblast invasion and cellular differentiation, which are important to the placenta and also to the success of the pregnancy. let-7a-5p expression in first trimester placental explants has been implicated in a reduction in cytotrophoblast proliferation. In our differential expression analysis we identified let-7a-5p as up-regulated in the 11-23 weeks’ gestation compared to the 6-10 weeks’ gestation group. Our detailed week-by-week expression data suggests an incremental increase across early to mid gestation which together with previous term data is consistent with let-7a-5p’s proposed role in regulation of placental growth by the reduction of cytotrophoblast proliferation [47]. The miR-34 family is known as a tumour suppressor miRNA family due to their synergistic effects with p53 [48]. Consistent with this, we identified up-regulation of all six miR-34 family members reflecting a reduction in cellular proliferation as the pregnancy progresses and placental growth trajectory begins to slow. Similarly, miR-378a-5p was previously found to enhance cell survival and promote trophoblast migration and invasion [49]. Our data showed significant down-regulation of the miR-34 family in 11-23 weeks’ gestation compared to the 6-10 weeks’ gestation placenta potentially linking down-regulation of these miRNAs with the reduced trophoblast migration as pregnancy progresses. miR-200 family members are also known to have an established role in placental function, in particular in the cellular transition from epithelial to mesenchymal (EMT) phenotype which is a prerequisite for extravillous trophoblast invasion of the maternal decidua. miR-200 family members play a well-known role in EMT through a feedback loop with the ZEB family of transcription factors. We identified seven members of the miR-200 family, six of which are down-regulated in the 11-23 weeks’ gestation group. The miR-200 family is known to be highly expressed in epithelial cell types [50] and thus its down-regulation is consistent with development of the vascularised mesenchymal core in chorionic villi in the second trimester.

### Proportional change in expression miRNA clusters play a role in placenta gene regulation during development

miRNA clusters are known to play an important role in the placenta, with these regions thought to have co-evolved and to act cooperatively to repress target genes [51]. Interestingly, while miRNA clusters are typically transcribed as a single polycistronic transcript, we identified large intra-sample variances in the expression levels of cluster members (Additional file 2, Figure S3) hinting at additional post transcriptional regulatory mechanisms. There are a number of miRNA clusters expressed either exclusively, or preferentially in placenta [18,52,53]. In particular, the C19MC, C14MC and miR-17∼92 clusters have been shown to regulate phenotypic and functional diversity in frequently used cell lines and isolated primary trophoblasts, and in the immune system, by which they may play a role in immune tolerance to paternal antigens [19,54,55].

C19MC is expressed almost exclusively in the placenta but also in certain tumors and undifferentiated cells [21,56]. C19MC is a large placenta specific imprinted cluster located on chromosome 19q13.41 and mono-allelically expressed from the paternally inherited chromosome [57]. This cluster has been difficult to study as its expression is both placenta and primate specific [57] and hence lacks an ortholog in mouse [22]. Importantly, C19MC members are expressed at lower than trophoblast levels in JEG-3 and JAR cells, two cell lines derived from choriocarcinoma [58], and are not expressed in the commonly used HTR8/SVneo cell line [53,56] highlighting the need for primary placental tissue in research. Consistent with our findings, previous research has shown that C19MC constitutes a large proportion of total miRNA transcripts (15% at term), with our data additionally showing a gestational decline from 27% at 6-10 weeks’ gestation to 23% at 11-23 weeks’ gestation [20,56]. In addition, there was a significant (FDR < 0.05) down-regulation in 34 C19MC cluster members in 11-23 weeks’ placenta compared to the 6-10 weeks’ gestation. Interestingly, a previous report showed that 46 of the 47 miRNAs from the C19MC cluster were up-regulated in third trimester villous trophoblasts compared to first trimester [19]. It is possible that this miRNA cluster exhibits a dynamic gestational expression pattern but perhaps more likely is the possibility that differences between studies can be explained by differences in expression between assessment of trophoblast versus whole chorionic villous tissue. The latter is more physiologically relevant and fits our question.

A recent miR-517a/c *in situ* hybridisation analysis localised expression from these C19MC miRs to cytotrophoblast and syncytiotrophoblast in chorionic villi, proximal cytotrophoblasts in anchoring villi, decreased expression in distal cytotrophoblasts with further reduction in extra-villous cytotrophoblasts. This finding highlights variable expression of the C19MC cluster in different placental cell types, and shows loss of expression of this cluster as trophoblasts differentiate to extravillous phenotypes post EMT [59]. Another report concurs showing higher expression of C19MC members in villous trophoblasts compared to extravillous trophoblasts suggestive of a role for the C19MC cluster miRNA in attenuation of EVT migration through the direct targeting of mRNA transcripts related to cellular movement [60]. Mouillet *et al*. (2015) suggested a role for C19MC in cellular differentiation or the maintenance of pluripotency [61]. Further investigation into the proportional change in expression from pregnancies complicated by placental pathology may help to identify miRNA species with biomarker potential in plasma. However, low expression levels in some maternal plasma samples have frustrated our efforts in this regard.

C14MC, located at the imprinted DLK1-DIO3 domain on human chromosome 14q32 is the largest known human miRNA cluster comprising 52 miRNA genes expressed from the maternally inherited chromosome [53]. The C14MC cluster is found exclusively in eutherian mammals and has been suggested to be essential to the evolution of this lineage, with expression in humans demonstrating a strong bias towards brain, placenta and some embryonic tissues [52]. Our analysis identified expression from 84 mature miRNAs in the C14MC cluster with the proportion of member transcripts increased in the 11-23 weeks’ gestation group compared to 6-10 weeks’ gestation. Previous studies investigating C14MC expression, in both whole villous tissue and in primary cytotrophoblasts, have reported a decrease in expression from first to third trimester. Using whole villous tissue, Gu *et al*. (2013) identified 11 miRNA from this cluster down-regulated in third compared to first trimester, and proposed a link between both C19MC and C14MC in immune suppression and innate/adaptive immune response in the mother. In primary cytotrophoblast, Morales-Prieto *et al*. (2012) identified 34 miRNA from this cluster which were also down-regulated in third compared to first trimester [53,62]. Whilst the discrepancy between our data and Morales-Prieto *et al*. (2013) can be explained by the use of different biological source (single cell type versus whole tissue), that between our data and Gu *et al*. (2013) is not so clear but may be due to differences in technology, the number of cluster members identified, or that our samples range up to 23 weeks’ gestation and do not include term. It is plausible that rather than a steady decline from early gestation to term, the expression of C14MC members increases across early gestation, before decreasing at term. Consistent with a recent report by Wommack *et al*. (2018) who profiled circulating miRNAs in maternal plasma and found an inverse relationship between placenta-specific clusters C19MC and C14MC we found this in our tissue samples.

Finally, in the analysis of placenta-associated clusters we investigated the expression of miR-17∼92 and paralogs miR-106a∼363 and miR-106b∼25, identifying 19 mature miRNAs. The proportion of total expression from miR-17∼92 members and paralogs are less than 1% per cluster, with all three clusters showing an overall decrease in the proportion of total expression across early to mid gestation. Using the miRNA sequences provided by Kumar *et al*. (2013), we determined the 5’ or 3’ origin of the transcripts by cross-checking the given sequences against miRBase v21[54,63], and were thus able to directly compare their microarray data with our sequencing data (detailed in Additional File 3: Table S11).

### Plasma miRNA sequencing offers a potential source of non-invasive biomarkers for detecting placenta health

In addition to establishing the placental miRNA profile across early to mid gestation and, given the potential for placental health biomarker discovery using our access to matched maternal plasma, we also aimed to analyse miRNA profiles in maternal plasma sampled in women at the same time as placenta. The identification of placenta derived, or pregnancy associated miRNA in plasma offers a potential window into minimally invasive monitoring of pregnancy progression.

Working with miRNA in plasma is known to be difficult due to potential inclusion of red blood cells or other cellular debris [64–66]. Red blood cell contamination may mask any signal we hoped to detect from the placenta as haemolysis may perturb miRNA species and count distributions due to overloading of red blood cell miRNAs [64,66,67]. In our study we see evidence that 11 of our samples were likely affected by haemolysis. This common difficulty with plasma data, led us to assess the detection of miRNAs of interest revealing that the 10 most abundant placental miRNAs were reflected by high abundance levels in matched plasma. Furthermore, these plasma miRNAs in our study show strong concordance with previous miRNA plasma studies [65,68]. Differential expression and clear gestational age gradient clustering pattern in placenta miRNAs between 6-10 and 11-23 weeks’ gestation were not particularly reflected by maternal plasma, indicating that plasma does not reflect the changes in the miRNA profile in placenta across early to mid gestation. miRNAs that were abundant in placenta were not always found to be abundant in plasma. In particular, miRNAs enriched in Pluripotent stem cell/embryonic stem cell were of low abundance in plasma compared to placenta. With the exception of placenta-specific miRNAs, cellular enrichment could be linked to various other systemic compartments. For example, five miRNAs have cellular enrichment in immune response cells, whilst another three miRNAs have enrichment in pluripotent and embryonic stem cells, and one miRNA is enriched in extra-embryonic stem cells which may originate from the fetal membranes or other extra-embryonic tissue such as chorionic villi. Of particular interest, miR-99a-5p enrichment includes enrichment in mast cells which are involved in inflammatory response and allergic reactions. We see an apparent increase in abundance of miR-99a-5p across early to mid gestation, and some additional increase in male-compared with female-bearing pregnancies which may be indicative of the increased maternal immune response to the male conceptus [69].

Complementary to the investigation of miRNA clusters in placenta, we also investigated the expression of three highly placenta-associated miRNA clusters and confirmed the abundance of these cluster miRNAs in plasma, with the proportion of cluster member expression revealing biomarker potential in plasma. One pathway for the transfer of placental molecules into the maternal circulation is via exosomes. It has previously been reported that members of the C19MC are the predominant miRNA species expressed in exosomes released from primary human trophoblasts [56]. We were able to confirm the presence of 39 C19MC, 48 C14MC and 20 miR-17∼92 miRNA in matched plasma, with their presence showing promise for the use of miRNA as biomarkers for placental health. Perhaps more importantly, week-by-week examination of miRNAs across early gestation, as described in this study, are key elements for developing pregnancy surveillance measures in the future, especially when combined with disease-specific profiles associated with preeclampsia, gestational diabetes and preterm birth. Our investigation here of miRNAs as biomarkers of placental, and by proxy pregnancy health have identified many areas for further investigation. With the implementation of machine learning algorithms, cell free DNA work and placenta specific analysis standards, much more can be expected from this field.

## Conclusion

Appropriate placental growth and development is essential for pregnancy success and miRNAs play roles in mediating these. We investigated placental and maternal plasma miRNAs using matched samples taken after elective termination of otherwise normal pregnancies and found 374 significantly (FDR <0.05) differentially expressed miRNA between 6-10 and 11-23 weeks’ gestation placenta.

The research conducted herein is the most comprehensive attempt to accurately profile the miRNA landscape of the human placenta, in part, because of the use of miRNA sequencing rather than microarray or qPCR. This dataset represents unprecedented access to a large number of human placenta samples from 6-23 weeks of gestation. Together these data are an important reference set for miRNA expression of early to mid gestation placental development and function, at time points rarely seen in previous placental sequencing work. These provide an important resource for the placental biology field. Further investigation using maternal plasma in early gestation samples from pregnancies with known outcomes will provide further insights into the search for miRNAs as biomarkers of pregnancy health and disease.

## Methods

### Sample collection

Placental chorionic villous tissue samples were obtained with informed, written consent from women undergoing elective terminations of otherwise healthy pregnancies. Samples of peripheral blood (6-9 mL) were collected from the same women into standard EDTA blood tubes at the time of termination and stored on ice until processed. Whole blood underwent centrifugation at 800 x g for 15 minutes at 4°C before plasma removal and then spun for a further 15 minutes. All samples were stored at −80°C until further processing. Prior to termination, accurate gestational age was determined using transvaginal ultrasonography. Termination samples were collected from the Pregnancy Advisory Centre (PAC), Woodville, South Australia. All placenta samples were assessed for gross morphology before inclusion in the study.

### RNA extraction and library preparation

All placenta tissue was collected post termination and processed as soon as possible after collection and placed into RNALater (Thermo Fisher) within 15 minutes. Total RNA was isolated using a modified protocol of the RNeasy mini plus kit (Qiagen, Hilden, Germany) whereby the RW1 buffer, a proprietary component of the RNeasy Kit that eliminates small RNAs, is replaced with 100% ethanol. The sample is then subjected to size fractionation for small RNAs before sequencing. For plasma, miRNA was isolated from 200 μL plasma samples using the QIAGEN miRNA serum/plasma kit (Qiagen, Hilden, Germany) according to the manufacturer’s instructions. All samples were stored at −80°C until further processing. miRNA real-time qPCR for assessment of haemolysis was conducted by QIAGEN Genomic Services (Qiagen, Hilden, Germany). 2ml RNA was reverse transcribed in 10 ml reactions using miRCURY LNA Kit (QIAGEN version5). Each RT was performed including an artificial RNA spike-in (UniSp6). cDNA was diluted 50 x and assayed in 10 ml PCR reactions according to the protocol for miRCURY LNA miRNA PCR; each miRNA was assayed once by qPCR using assays for miR-23a, miR-30c, miR-103, miR-142-3p, and miR-451. In addition to these miRNA assays, the RNA spike-ins were assayed. The amplification was performed in a LightCycler® 480 Real-Time PCR System (Roche) in 384 well plates. The amplification curves were analysed using in-house software, both for determination of the Cq (by the 2^nd^ derivative method) and for melting curve analysis.

All placenta samples were genotyped for fetal sex using high resolution melt curve analysis of the gene that encodes amelogenin. Amelogenin is found on both the X (AMELX) and Y (AMELY) chromosomes, the X allele features a 3 bp deletion in exon 3 allowing the identification of samples with only X chromosomes (female) or X and Y chromosomes (male) prior to sequencing. Forward primer 5’-CCCTGGGCTCTGTAAAGAATAGTG-3’, reverse primer 5’- ATCAGAGCTTAAACTGGGAAGCTG-3. qPCR was performed using SSOFast EvaGreen Supermix (Bio-Rad, CA), primers at 250 nM final concentration, 5 ng of DNA per reaction, on a Bio-Rad CFX384 Real-Time PCR System. Cycling conditions: initial denaturation 98°C 30s, 40 cycles of 98°C for 5s and 60°C for 5s. High resolution melt curve analysis was performed from 65°C to 85°C with a 0.2°C increment every 10s. Melt curve between 65°C and 70°C was analysed using Bio-Rad Precision melt software (Bio-Rad, CA) to identify sex genotypes. Library preparation and sequencing was performed by Qiagen (Valencia, CA) using the QIAseq miRNA Library Kit and QIAseq miRNA 48 Index IL kits as per manufacturer’s instructions. Amplified cDNA libraries underwent single-end sequencing by synthesis (Illumina 1.9).

### High-throughput sequencing analysis

Alignment and analysis of miRNA sequencing data were processed from raw FASTQ files using the *bcbio-nextgen* pipeline [70]. Briefly, adapter detection and trimming were performed using Atropos [71]. Alignment was performed using STAR [72] using the human reference genome build GRCh37 [73] (https://www.ncbi.nlm.nih.gov/grc). Quality control metrics were assessed using FastQC [74] (http://www.bioinformatics.babraham.ac.uk/projects/fastqc/) to check for per base sequence quality, sequence length distribution and duplication levels, and summarised using multiQC [75]. miRNA were clustered and collapsed using SeqCluster [76], and individual miRNAs detected using SeqBuster [70]. Annotation was performed using the miraligner [70] with miRBase version 21.0 [15,63] (http://www.mirbase.org) as the reference database.

### Differential expression analysis

All profile and expression analyses were conducted in the R statistical environment (v.3.3.2), using the *edgeR* (v.3.16.5) and *limma [77]* (v.3.30.11) R/Bioconductor packages. *edgeR [78]* was used to filter miRNA with low expression and normalise for library composition bias. Batch effects, introduced by sequencing samples in multiple sequencing runs, were evident after normalisation and corrected using a *limma* batch correction method [77]. All samples were then normalised using the Trimmed Mean of M values (TMM).

Sample-weights and log transformation was performed using *limma* package [79] with the voom function used to estimate the mean-variance relationship between individual observations and then applied to the normalised log-counts data. Differential expression analysis including, moderated F-statistic evaluation, adjusted p-value estimation and Log Fold Change (LFC) analysis were performed using a moderated t-test [80] with Benjamini-Hochberg (BH) multiple hypothesis test corrections [81]. After adjustment, expression of miRNAs was considered significantly different at FDR ≤ 0.05. The workflow, source code and input files associated with this research available at (https://github.com/mxhp75/earlyPlacentamiRNA_SeqProfile.git).

## Data Availability

The dataset(s) supporting the conclusion of this article are available in NCBI's Gene Expression Omnibus [82] and are accessible through GEO Series accession number GSE151362

https://www.ncbi.nlm.nih.gov/geo/query/acc.cgi?acc=GSE151362

https://github.com/mxhp75/earlyPlacentamiRNA_SeqProfile.git

## Declarations

### Ethics approval and consent to participate

Ethics approval for the collection of placenta tissue and blood from women undergoing elective pregnancy termination between 6–23 weeks’ gestation was provided under HREC/16/TQEH/33, by The Queen Elizabeth Hospital Human Research Ethics Committee (TQEH/LMH/MH).

### Consent for publication

#### Availability of data and materials

The dataset(s) supporting the conclusion of this article are available in NCBI’s Gene Expression Omnibus [82] and are accessible through GEO Series accession number GSE151362 (https://www.ncbi.nlm.nih.gov/geo/query/acc.cgi?acc=GSE151362).

#### Funding

This research is supported by NIH NICHD R01 HD089685-01 Maternal molecular profiles reflect placental function and development across gestation PI Roberts, an Australian Government Research Training Program (RTP) Scholarship awarded to MDS, a National Health and Medical Research Council Investigator Grant (GNT1174971) awarded to CTR and a Matthew Flinders Professorial Fellowship awarded to CTR and funded by Flinders University. JB is supported by the James & Diana Ramsay Foundation.

### Authors contributions

CTR created the concept. MS, CTR, JB and TBM conceived and developed experimental plans. TJK, DMcA and DMcC performed experiments. MS and JB analysed the sequencing data with help from KB and QW. Manuscript written by MS, JB and CTR with help from TBM and KP. All authors read and approved the final version of the manuscript.

## Acknowledgements

We wish to acknowledge the generosity of the women who donated their placenta for our research. Without them, this research would not be possible. We also acknowledge valuable input from QIAGEN Genomic Services.

## Author information

^1^Robinson Research Institute, ^2^Adelaide Medical School, ^3^Bioinformatics Hub, ^4^The University of South Australia, ^5^School of Agriculture Food and Wine; University of Adelaide, ^6^South Australian Health & Medical Research Institute (SAHMRI), ^7^Flinders Health and Medical Research Institute, Flinders University

## Additional file

### Additional_File_1.xlxs

**Table S1**. Read statistics for 96 placenta samples miRNA-Seq data. **Table S2**. miRNA expression data from 86 placenta chorionic villous samples. Expression values calculated as log2 CPM across 86 samples passing filters (all_mean), across samples of 6-10 weeks’ gestation (≤10_mean) and for samples 11-23 weeks’ gestation (>10_mean). Relative ranks for mean expression is also given as all_rank, ≤10_rank and >10_rank respectively. **Table S3**. Principal component analysis results showing the contribution of each miRNA across PC1 and PC2. **Table S4**. Differential expression analysis of data from 86 placenta chorionic villous samples identified 374 significant (FDR <0.05) different miRNA between 6-10 weeks’ and 11-23 weeks’ gestation groups. **Table 5**. Differential expression analysis of data from 86 placenta chorionic villous samples revealed 34 C19MC cluster miRNAs down-regulated in the 6-10 weeks’ compared to the 11-23 weeks’ gestation groups. **Table S6**. Differential expression analysis of data from 86 placenta chorionic villous samples revealed 56 C14MC cluster miRNAs with significant (FDR <0.05) differential expression between the 6-10 weeks’ compared to the 11-23 weeks’ gestation. **Table S7**. Differential expression analysis of data from 86 placenta chorionic villous samples revealed 17 miR∼17-92 cluster miRNAs with significant (FDR <0.05) differential expression between the 6-10 weeks’ and 11-23 weeks’ gestation groups.

### Additional_File_2.docx

**Figure S1**. Using principal component analyses of the miRNA-Seq data from 94 placenta samples, we identified 8 samples (named) that segregated from the remaining 86 samples. This separation on PC1 is most likely due to the presence of surrounding non-villous tissue. **Figure S2**. The PCA plot of batch corrected data shows no gestational age gradient for maternal plasma from 6-23 weeks’ gestation. The four samples in the upper right quadrant were sequenced in the same sequencing run and may represent a technical artefact in line with previous difficulties in plasma sequencing but have not been removed from further analysis. **Figure S3**. Heatmap of C19MC cluster members plotted from 5’ (left) to 3’ (right), and from early (top) to later (bottom) gestation clearly identifies different expression levels for the polycistronic members of this cluster.

### Additional_File_3.xlxs

**Table S8**. Read statistics for 96 plasma samples miRNA-Seq data. **Table S9**. miRNA abundance data from 96 maternal plasma samples. Expression values calculated as log2 CPM across 96 samples. miRNA ranked 1:790 by abundance. **Table S10**. Differential expression analysis of data from 86 placenta chorionic villous samples using gestational age (6-23 weeks’) as a continuous variable. **Table S11**. We compared our T1 vs T2 differential expression data with those of Kumar et al. (2013) who compared expression of miR-17∼92, and its paralogs miR-106a∼363 and miR-106b∼25 clusters and identified a down-regulation of cluster members post syncytialisation.

