## Additional file 2 for "Large-scale transcriptome-wide profiling of microRNAs in human placenta and maternal plasma at early to mid gestation"

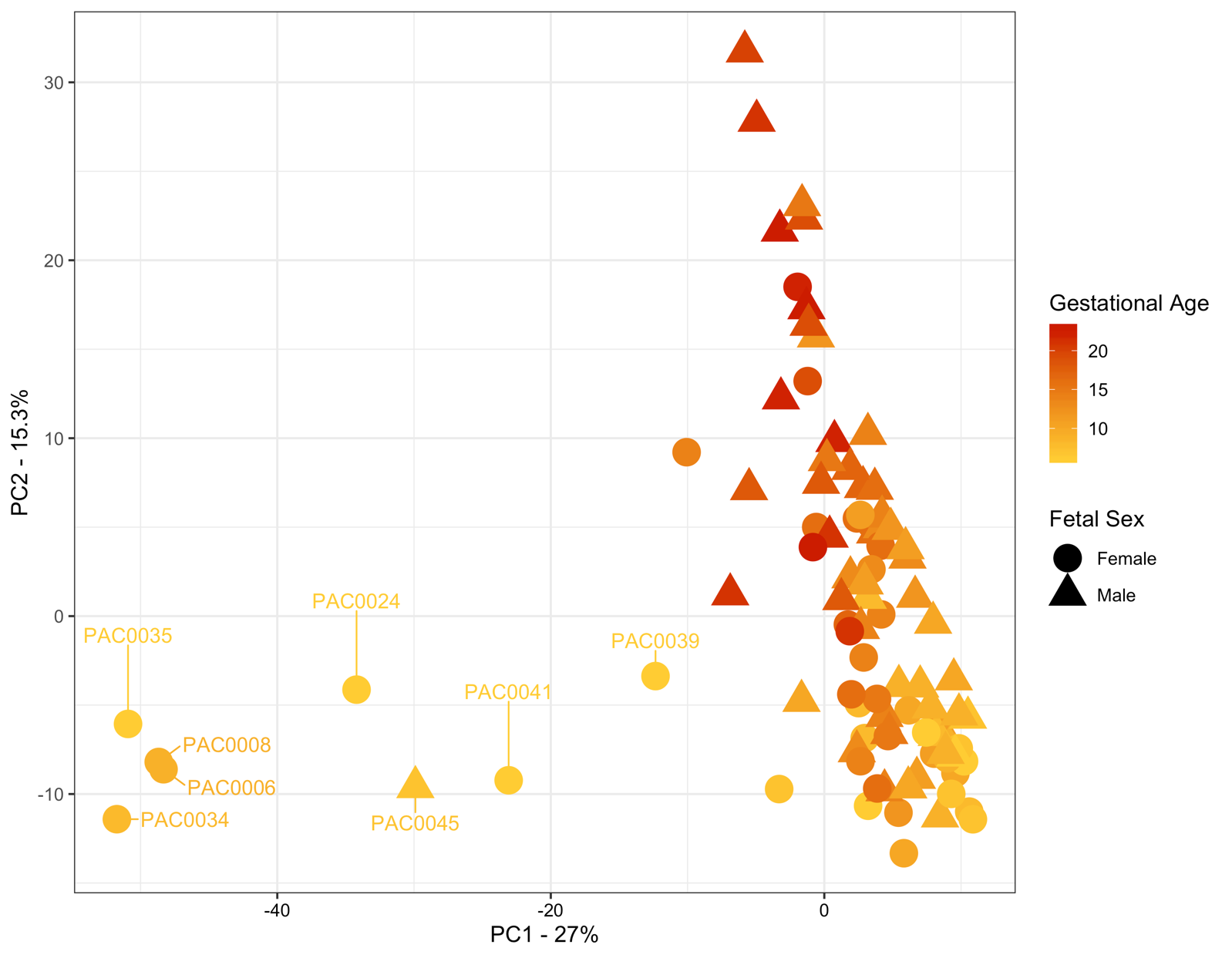

**Additional File 2: Figure S1.** Using principal component analyses of the miRNA-Seq data from 94 placenta samples, we identified 8 samples (named) that segregated from the remaining 86 samples. This separation on PC1 is most likely due to the presence of surrounding non-villous tissue.

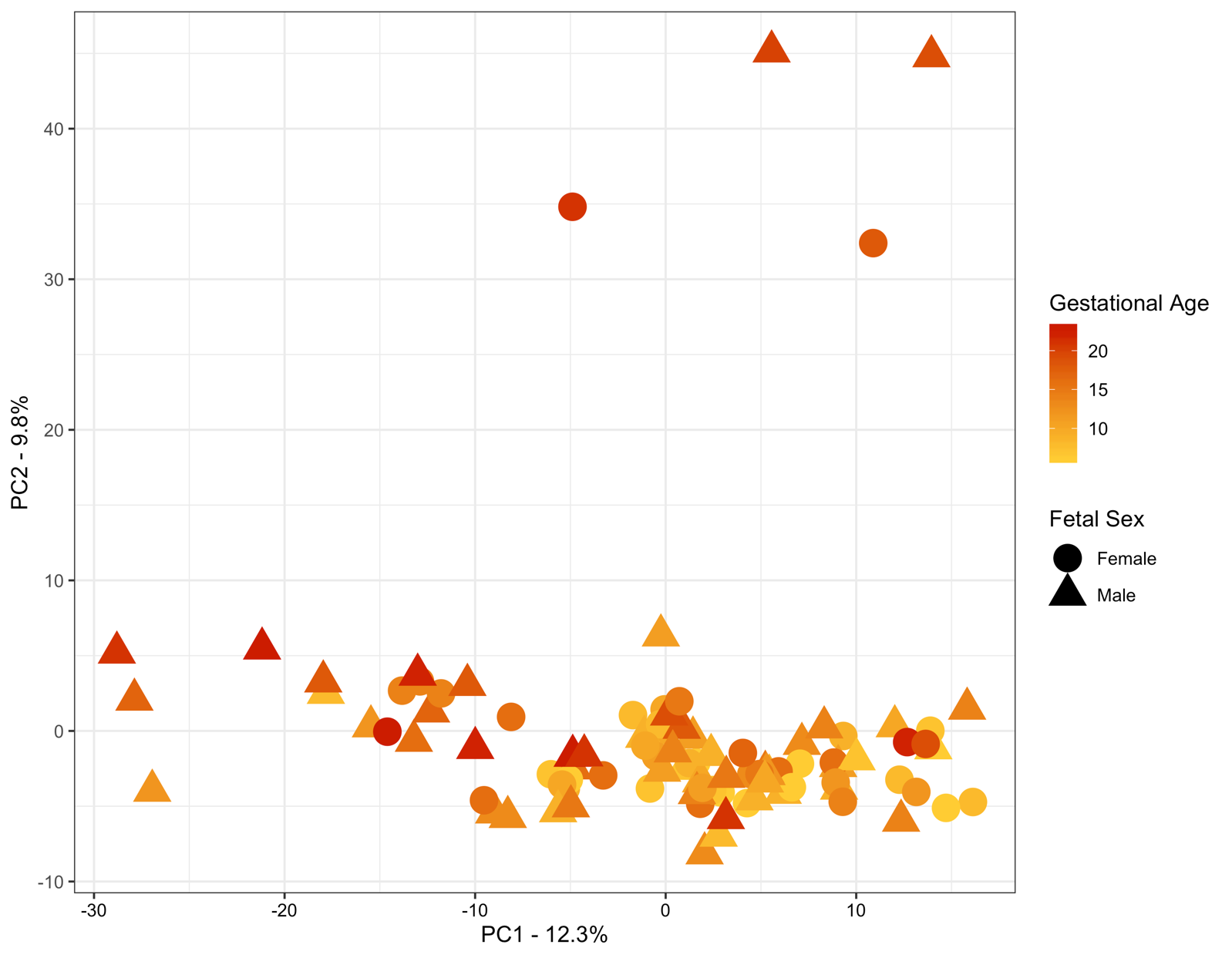

**Additional File 2: Figure S2.** The PCA plot of batch corrected data shows no gestational age gradient for maternal plasma from 6-23 weeks’ gestation. The four samples in the upper right quadrant were sequenced in the same sequencing run and may represent a technical artefact in line with previous difficulties in plasma sequencing but have not been removed from further analysis.

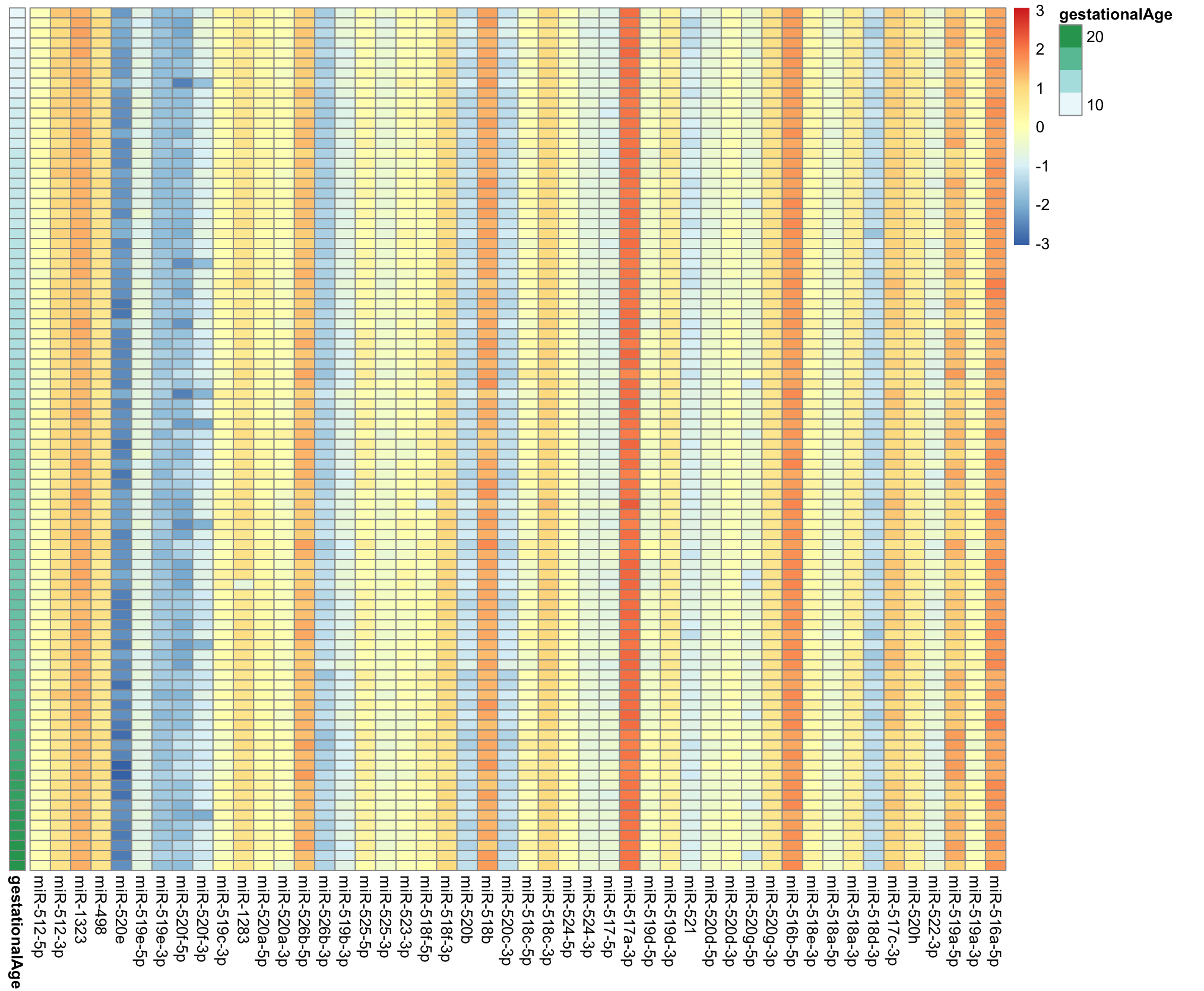

**Additional File 2: Figure S3.** Heatmap of C19MC cluster members plotted from 5` (left) to 3` (right), and from early (top) to later (bottom) gestation clearly identifies different expression levels for the polycistronic members of this cluster.
